# Novel MAPK/AKT-impairing germline *NRAS* variant identified in a melanoma-prone family

**DOI:** 10.1101/2021.04.20.21254596

**Authors:** Kevin M. Brown, Mai Xu, Michael Sargen, Hyunbum Jang, Mingzhen Zhang, Tongwu Zhang, Bin Zhu, Kristie Jones, Jung Kim, Laura Mendoza, Nicholas K. Hayward, Margaret A. Tucker, Alisa M. Goldstein, Xiaohong Rose Yang, Douglas R. Stewart, Belynda Hicks, Dario Consonni, Angela C. Pesatori, Maria Concetta Fargnoli, Ketty Peris, Alex Stratigos, Chiara Menin, Paola Ghiorzo, Susana Puig, Eduardo Nagore, MelaNostrum Consortium, Thorkell Andresson, Ruth Nussinov, Donato Calista, Maria Teresa Landi

## Abstract

While several high-penetrance melanoma risk genes are known, variation in these genes fail to explain melanoma susceptibility in a large proportion of high-risk families. As part of a melanoma family sequencing study, including 435 families from Mediterranean populations, we identified a novel *NRAS* variant (c.170A>C, p.D57A) in a melanoma-prone family. This variant is absent in exomes in gnomAD, ESP, UKBiobank, and the 1000 Genomes Project, as well as in 11 273 Mediterranean individuals and 109 melanoma-prone families from the US and Australia. This variant occurs in the GTP-binding pocket of NRAS. Differently from other RAS activating alterations, NRAS D57A expression is unable to activate MAPK-pathway both constitutively and after stimulation but enhances EGF-induced PI3K-pathway signaling in serum starved conditions *in vitro*. Consistent with *in vitro* data demonstrating that NRAS D57A does not enrich GTP binding, molecular modeling suggests that the D57A substitution would be expected to impair Mg2+ binding and decrease nucleotide-binding and GTPase activity of NRAS. While we cannot firmly establish NRAS c.170A>C (p.D57A) as a melanoma susceptibility variant, further investigation of *NRAS* as a familial melanoma gene is warranted.

## Introduction

Melanoma incidence has been increasing in Australia, Europe, and the United States for several decades. In 2018, there were 290 000 cases of melanoma worldwide, and approximately 5-10% of cases occur in individuals with a family history of melanoma (1-4). Multiple melanoma-predisposition genes (*CDKN2A, CDK4, BAP1, POT1, TERT, ACD, TERF2IP*) explain melanoma susceptibility in 20-30% of melanoma-prone families, suggesting that inherited risk for most families results from multiple moderate to low penetrance susceptibility genes or yet unidentified high-penetrance susceptibility genes (5).

Somatic *NRAS* mutations are identified in approximately 20% of primary melanomas and occur in tumors that are *BRAF* wild type (6). In contrast, germline variants of *NRAS* and other genes of the *RAS* pathway predispose to syndromes including Noonan Syndrome (7). In this report we describe a family with a novel germline variant of *NRAS* (c.170A>C, p.D57A) in which affected carriers developed melanoma and autoimmunity.

## Results

### Family clinical history

The proband of the family (**Fig 1A, S1 Fig**) was diagnosed with an invasive cutaneous melanoma of his right arm at age 35-39, which was successfully treated with wide local excision. One year later, he was diagnosed with Sjogren’s syndrome after serologies were ordered for chronic xerophthalmia. His symptoms were successfully controlled with artificial tears and did not require systemic treatment. At age 60-64, the patient was diagnosed with an oncocytoma in his right kidney. Prior to this diagnosis, the patient denied any history of pneumothorax or prior skin biopsies demonstrating fibrofolliculomas/trichodiscomas to suggest a diagnosis of Birt– Hogg–Dubé (BHD) syndrome (OMIM: 135150). On clinical exam, the patient also did not exhibit any white or pink papules on the face that would be consistent with fibrofolliculomas/trichodiscomas.

**Fig 1.**
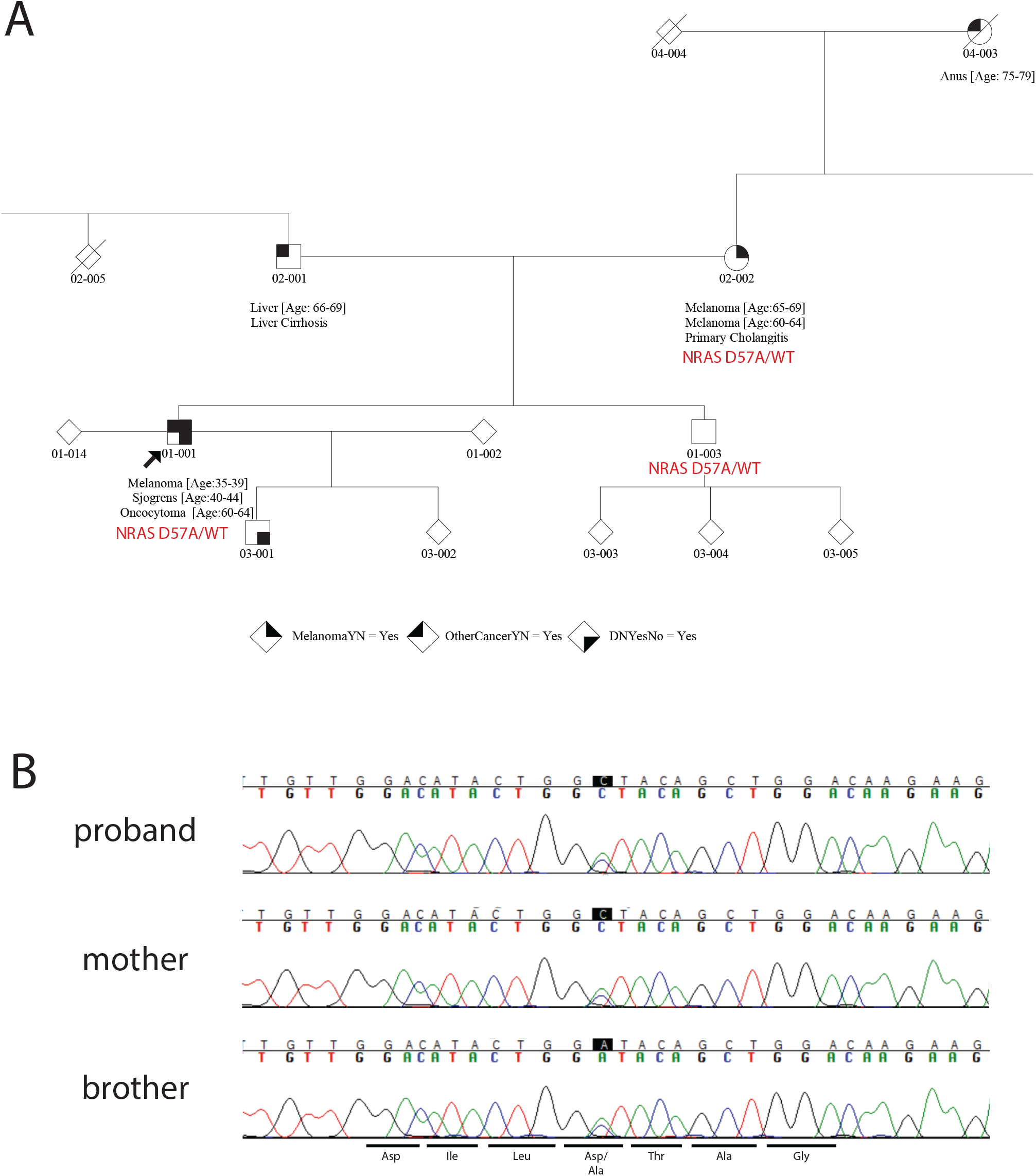
Pedigree of melanoma family carrying *NRAS* c.170A>C (p.D57A). (A) Pedigree of the immediate family harboring *NRAS* c.170A>C (p.D57A) annotated with sequencing and genotyping results. Individuals diagnosed with melanoma, other cancers, and dysplastic nevi are annotated, and the proband is denoted with an arrow. (B) Sanger sequencing traces of c.170A>C/p.D57A variant for familial carriers.

The proband’s mother had two melanomas, with the first diagnosis on the back at age 60-64 and the second diagnosis on the face at age 65-69. She was also diagnosed with primary biliary cholangitis as a young adult. The proband’s maternal grandmother had anal cancer (reported by the proband as possibly mucosal melanoma, but the diagnosis is unconfirmed), but no history of autoimmune disease. The proband’s father was diagnosed with liver cancer at age 65-69, which was thought to be secondary to chronic HCV infection. The proband has two healthy children, both without any history of autoimmune disease, melanoma, or other cancers; his son was diagnosed with clinically dysplastic nevi (as was the proband). The family history was negative for other malignancies. Syndromic features of RASopathies, including facial dysmorphism, cardiac abnormalities, and skin findings (lentigines, hemangiomas), were not present in the family.

### Genetics

As part of a larger whole-exome sequencing (WES) study of 435 high-risk melanoma families [675 affected individuals or obligate carriers sequenced] of Mediterranean ancestry (8), germline sequencing of the proband and his mother revealed the presence of a shared novel variant in *NRAS* [g.chr1:115256541T>G, NM_002524.4 c.170A>C, NP_002515.1 p.D57A]; both were mutation-negative for known melanoma susceptibility genes (*CDKN2A, CDK4, BAP1, POT1, TERT, ACD, TERF2IP, and MITF* E318K); the proband did not carry any pathogenic variants in genes associated with oncocytoma (*FLCN, TSC1*, or *TSC2*) or monogenic autoimmune syndromes. Sanger sequencing confirmed this variant in the proband, proband’s mother, and proband’s unaffected brother (**Fig 1A and 1B**), with all individuals being heterozygous. No other rare or novel *NRAS* variants were found in other melanoma families. This variant is classified as likely pathogenic by InterVar (applying ClinGen RASopathy recommendation) (9) and predicted deleterious by multiple *in-silico* prediction tools (REVEL=0.924, CADD=24.3, metaSVM=damaging). The variant has not been observed in ClinVar or HGMD. In addition, similar to somatic activating mutations found in melanomas and nevi (codons 12, 13, and 61) (10, 11), or alternatively in the germline of families with a group of syndromic conditions broadly termed rasopathies (codons 12, 13, 50, 58, and 60) (12), D57 is located in the RAS GTP-binding pocket.

This variant has not been reported in gnomAD, ESP, UKBiobank, and 1000 Genomes Project other germline codon 57 variants appear to be exceptionally rare, with only a single non-synonymous codon 57 allele (1:g.115256542C>T, c.169G>A, p.D57N) found in gnomAD (1/113 658 non-Finnish European alleles). While confirming the presence of the variant in DNA from the proband and his mother (used as positive controls), TaqMan genotyping of samples from the MelaNostrum Consortium (13, 14) (**S1 Table**) found this alteration to be altogether absent in 5 184 alleles of the same national ancestry as the proband (including both melanoma cases and healthy controls; hundreds of individuals were from the same area of the country from which the family comes), as well as 12 958 additional alleles of Mediterranean ancestry (of different countries as the proband). We also examined additional WES data generated from participants of the EAGLE (15) study, with this variant absent in 4 404 alleles (1 371 lung cancer cases and 831 healthy controls) as well in 308 melanoma cases from 109 high-risk melanoma families from the US and Australia.

### *In vitro* characterization of NRAS D57A

The D57 residue of NRAS is highly conserved in RAS orthologs and paralogs and located within a guanine nucleotide-binding pocket (16). Nearby germline variants T50I and G60E of *NRAS* have been reported in patients with Noonan Syndrome (7), and *in vitro* characterization showed that these alterations enhanced stimulus-dependent MAPK activation, similar to several somatically activating mutations previously identified in melanomas (G12V, Q61R). To explore the possible functional consequences of the *NRAS* c.170A>C (D57A) variant, we expressed it in COS7 cells, and its effect on MAPK and PI3K signaling pathway was compared with both wild-type NRAS and the oncogenic NRAS G12V mutation. While NRAS G12V stimulated constitutive activation of both ERK and AKT, as well as EGF-mediated activation of ERK and AKT under serum-starved conditions, NRAS D57A specifically enhanced AKT but not ERK phosphorylation only upon treatment of human EGF under serum starvation (**Fig 2A**). Furthermore, enhanced AKT activation via NRAS D57A expression disappears with increased dose of EGF, suggesting that this variant may function under sub-optimal growth conditions (**Fig 3**). In addition, D57A NRAS appeared to be deficient in activation of MAPK signaling even in the presence of serum. As D57 is part of the guanine nucleotide-binding pocket and NRAS function is regulated by its nucleotide binding status, we pulled down GTP-bound NRAS with RAF1 RAS-binding domain-conjugated agarose beads. Unlike WT NRAS and NRAS G12V, D57A NRAS was not enriched in the GTP-bound state (**Fig 2B**), seemingly contradictory to enhanced AKT activation upon EGF treatment but consistent with the overall observed lack of MAPK activation. Of note, we also consistently observed less expression of D57A NRAS than either wild-type NRAS or G12V NRAS, and the NRAS D57A band migrates faster through gels compared to its WT or G12V counterpart, perhaps suggesting altered protein stability and/or post-translational modification.

**Fig 2.**
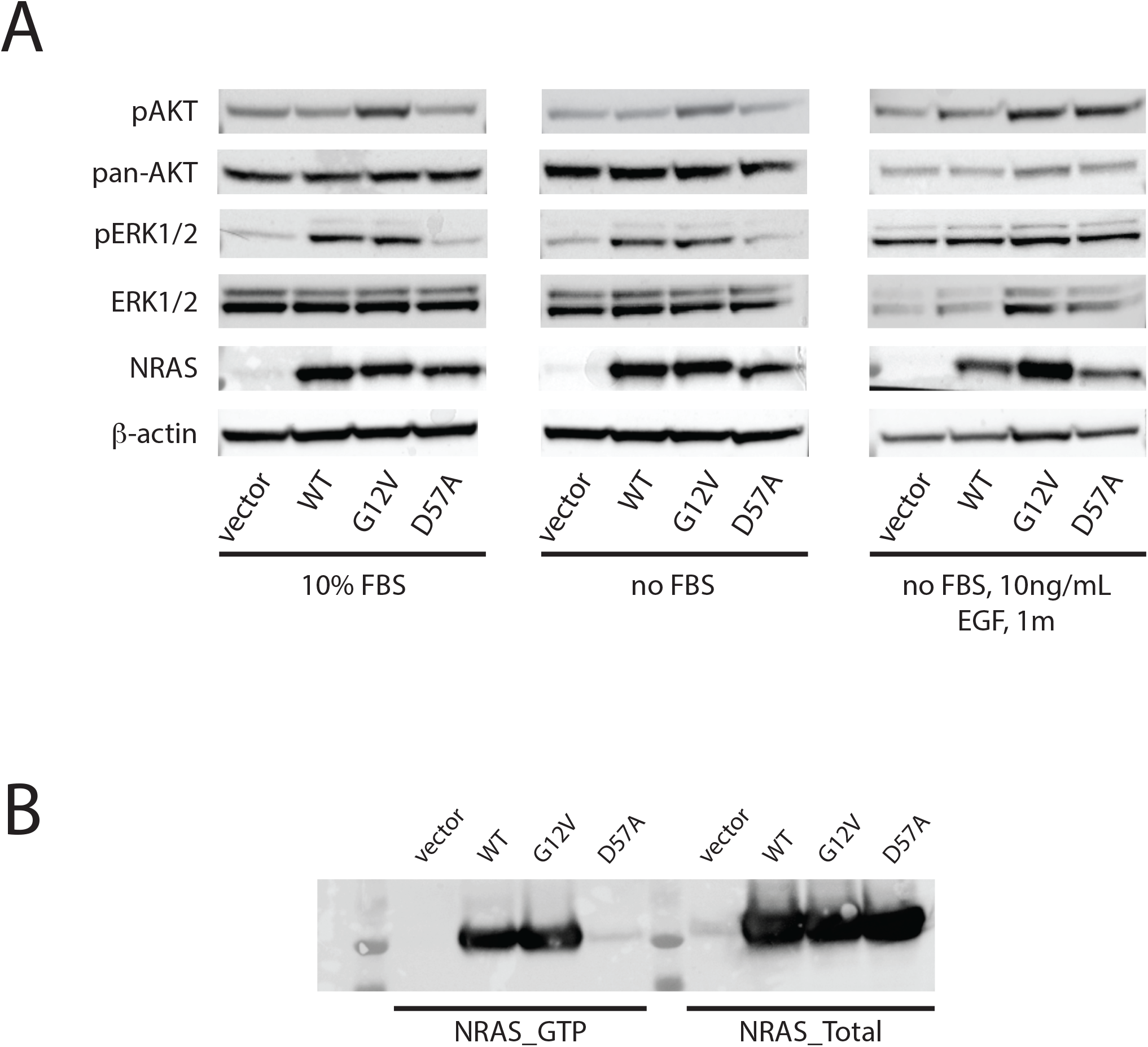
NRAS D57A increases AKT signaling after EGF stimulation in serum-starved cells. (A) wild-type NRAS, as well as G12V and D57A NRAS variant were transiently transfected into COS7 cells and were switched to serum-free medium at 24hrs after transfection, followed by EGF treatment (10 ng/mL, 1m) after one day of serum starvation. Pan- and phospho-ERK and - AKT were assessed by western blot. (B) WT NRAS, as well as G12V and D57A NRAS mutations were transiently transfected in 293FT cells, and at day 2 after transfection, whole cell extract was taken to pull down GTP-bound NRAS by RAF1_RBD conjugated agarose beads, followed by western blotting. Left Panel: NRAS blot after pull-down; Right Panel: NRAS blot before pull-down.

**Fig 3.**
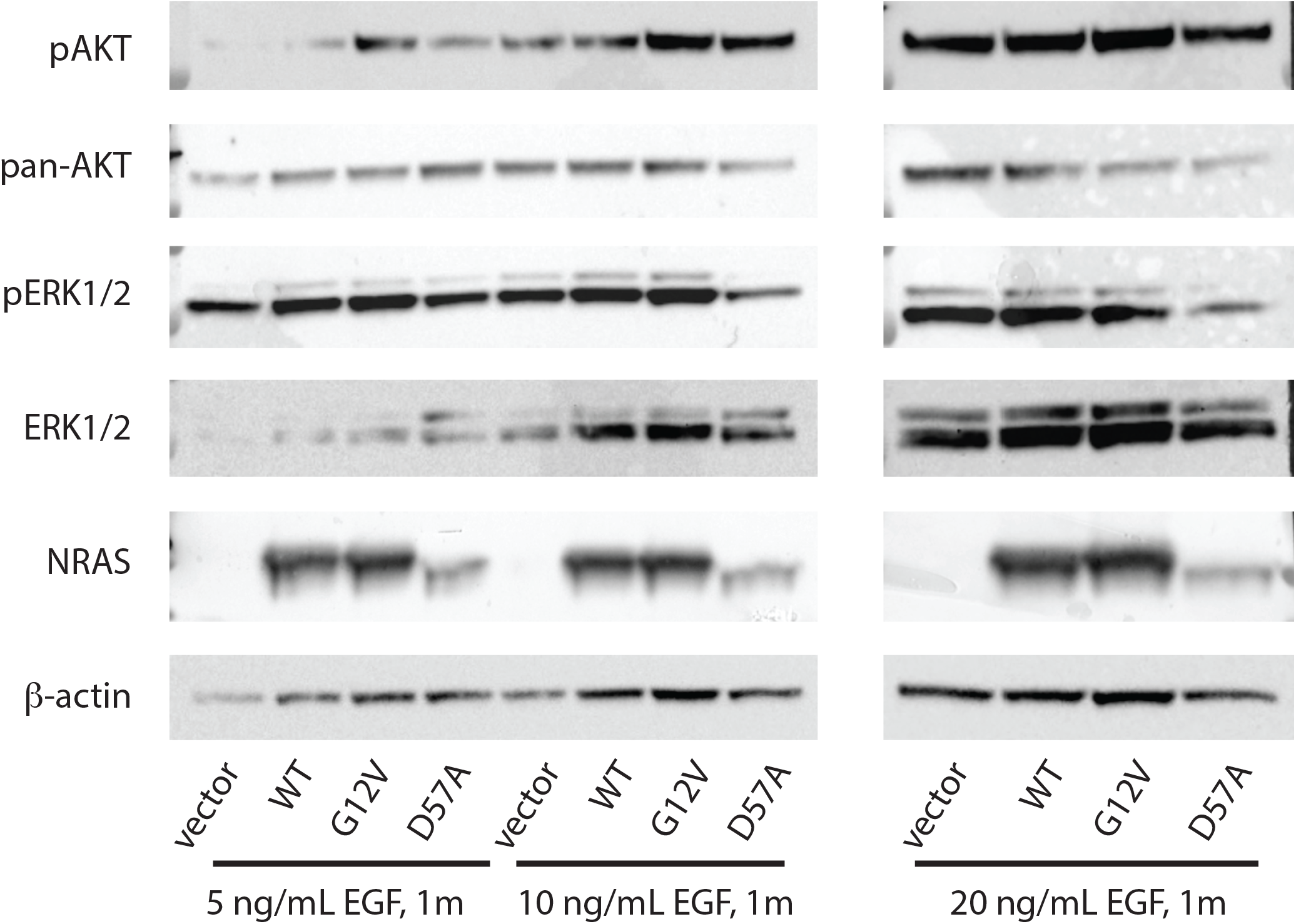
NRAS^D57A^ effect on AKT signaling depends on EGF dose. Constructs expressing wild-type NRAS, as well as G12V and D57A NRAS mutations were transiently transfected into COS7 cells and were switched to serum-free medium at 24hrs after transfection, followed by EGF treatment (5, 10, and 20 ng/mL for 1 minute) after one day of serum starvation. Pan- and phospho-ERK and -AKT were assessed by western blot.

### Molecular modeling of NRAS D57A protein

*In vitro* data are consistent with molecular modeling of this variant. As most activating RAS mutations activate both MAPK and AKT signaling pathways, NRAS D57A appears to be a unique RAS variant that specifically activates the AKT pathway under low stimulus conditions. Visualization of the RAS nucleotide-binding pocket by neutron crystallography indicates that protonation of D57 will impair Mg^2+^ retention, thus decrease nucleotide-binding activity and GTPase activity of RAS, making it less stable in structure (16). The D57A variant identified in the melanoma family would be expected to have the same effect as protonation of D57 and as a result, this variant may be predicted to affect its catalytic activity as well as protein stability.

To further characterize the biological effects of the D57A alteration, we used molecular modeling to analyze the molecular behavior of NRAS D57A. We performed the 500 ns simulations for both wild-type NRAS and NRAS D57A in the explicit solvent. D57 contributed to the coordination of Mg2+ in NRAS, with a distance of ∼4.5Å. Upon D57A mutation, the distance of A57 to Mg2+ is predicted to increase to ∼7.0Å (**Fig 4A-4C**). This is consistent with the molecular findings where D57A NRAS protein was not enriched for its GTP-bound state. In NRAS, D57 interacts with T35 (switch I region) via hydrogen bonds and coordinates the Mg2+ ion along with S17. In simulations, the D57A variant disrupted the local residue contacts, increasing the side chain distances (**Fig 4D-4E**). These changes in the local residue contacts by the D57A mutation induce a conformational change of switch I region. The side chain of Y32 reorients towards the GTP’s γ-phosphate, with the distance decreased (**Fig 4F**). Notably, D57A NRAS affects the PI3K/AKT pathway but is seemingly deficient in MAPK-pathway activation *in vitro* relative to wild-type NRAS (**Fig 2A**). A previous study investigating the effects of man-made mutations to the GTP-binding pocket of HRAS observed that D57A decreases binding to CRAF *in vitro* (17). RAS interacts with the RBD of RAF in the MAPK pathway mainly through the switch I region (18, 19), where both the switch I and II regions mediate the RAS interactions with PI3K in the PI3K/AKT pathway (20). The conformational change of switch I upon D57A mutation is expected to affect the RAF interactions. In sum, the D57A substitution is predicted to impair Mg^2+^ retention, decrease nucleotide-binding and GTPase activity of RAS, and likely making it less stable in structure (16), suggesting NRAS D57A is a functional variant enhancing AKT activation under some conditions, as well as potentially altering nucleotide binding activity and protein structure.

**Fig 4.**
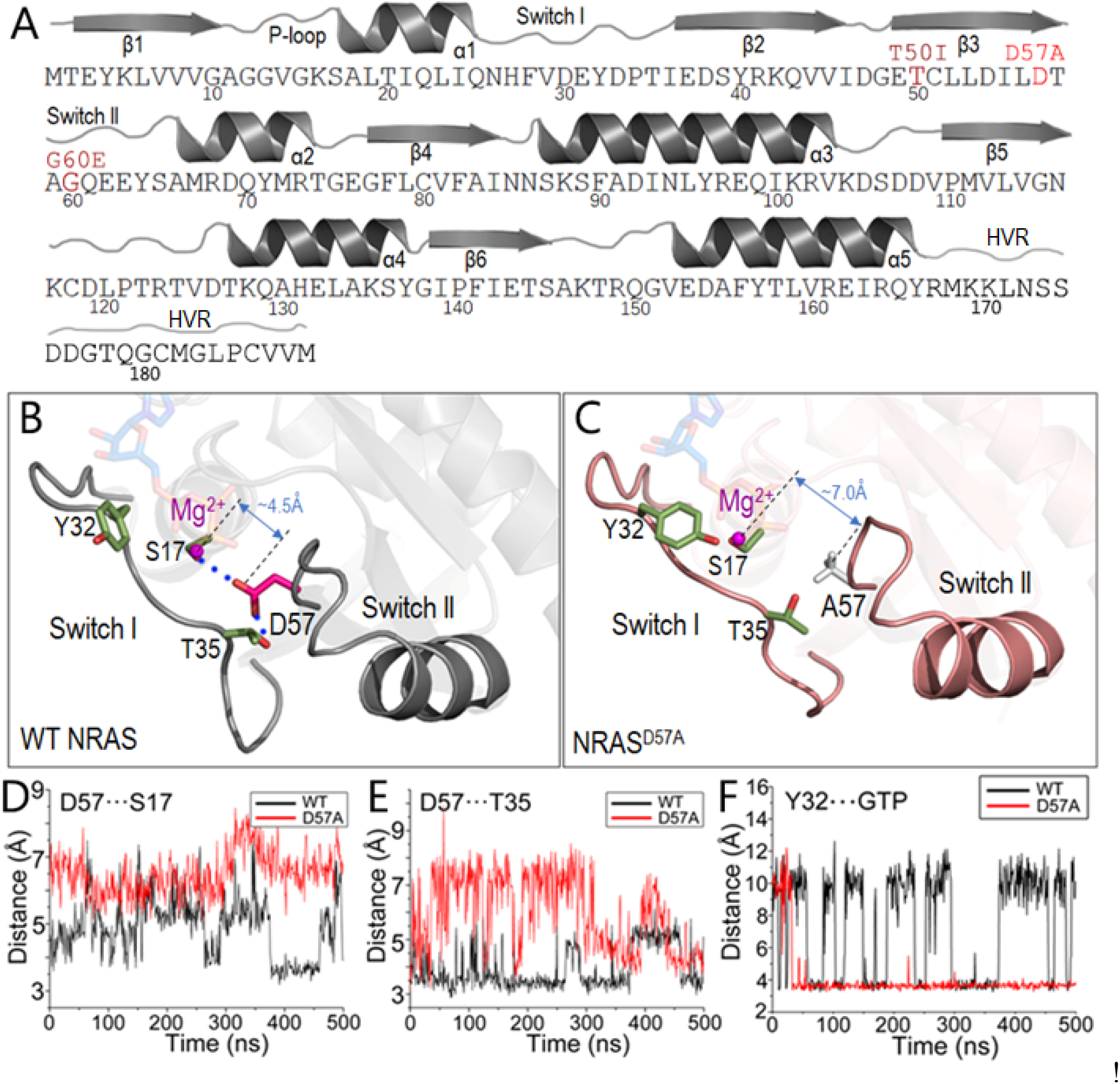
Structural insight into *NRAS*^D57A^. (A) The sequence of NRAS with select mutations highlighted (T50I and G60E for Noonan Syndrome, and D57A for melanoma). (B) The representative snapshots of wild-type *NRAS* and (C) *NRAS*^D57A^ from molecular dynamics (MD) simulations. Upon D57A mutation, the interactions of D57 with (D) S17 and (E) T35 are disrupted, with the distances increased. The side chain of Y32 reorients towards GTP with (F) the distance decreased, suggesting a conformational change of switch I region upon D57A mutation.

## Discussion

As part of a large melanoma family exome-sequencing study, we identified a novel germline variant of *NRAS* (c.170A>C /p.D57A) in two melanoma cases from a melanoma family. This variant has not been identified in any population to date (cataloged by gnomAD, 1000 Genomes Project, ESP, and UKBiobank), we did not observe it in >22 000 Mediterranean alleles (including >9 500 alleles from the same country as the proband) and in 109 melanoma-prone families from the US and Australia, and germline alteration of *NRAS* codon 57 appears to be exceedingly rare, with only one single mutant allele (D57N) cataloged by gnomAD. Further, multiple *in-silico* prediction tools predict this variant to be pathogenic.

*In vitro* assays and molecular modeling suggest that this variant, located within the GTP-binding pocket of NRAS where other recurrent functional germline and somatic variants cluster, clearly alters NRAS function. Notably, while well-characterized germline and somatic variants of RAS proteins most often result in activation of either both PI3K and MAPK, *in vitro* assays demonstrate D57A did not activate MAPK in the presence of serum and enhanced only AKT phosphorylation via EGF under serum starvation, making this subtly-activating variation unique. Consistent with these findings, the corresponding mutation in HRAS has been previously shown to result in decreased binding of HRAS to CRAF (17) *in vitro*. Our *in vitro* assays additionally suggest D57A may result in altered protein stability and or post-translational modification, and molecular modeling suggests this mutation is likely to alter Mg^2+^ retention and decrease nucleotide-binding activity and GTPase activity.

Given the small size of this family, it is difficult to assess the impact of this alteration on melanoma and/or autoimmune syndrome risk, as (1) we do not observe *NRAS* variation in other melanoma families, (2) observed cosegregation with melanoma may be due to chance, (3) we observe this variant in the proband’s sibling who has been unaffected to date, and (4) no germline material is available from other members of this family diagnosed with melanoma or with autoimmune symptoms, nor are tissues from the proband or his mother.

In conclusion, we identified a novel, functionally significant germline variant of *NRAS* in a melanoma-prone family. While a direct association with melanoma risk cannot be firmly established from this observation, investigation of *NRAS* variants potentially harbored by other melanoma-prone families including those drawn from other populations, is warranted. The proband was also diagnosed with both Sjogren’s syndrome and oncocytoma; further investigation of families with autoimmune diseases and/or kidney tumors may shed additional light on whether this mutation may influence risk of these diseases.

## Materials and Methods

### Whole exome sequencing of melanoma families

675 melanoma cases or obligate carriers from 435 multi-case *CDKN2A/CDK4* mutation-negative melanoma families (2-5 affected relatives) of Mediterranean ancestry from the MelaNostrum Consortium (https://dceg.cancer.gov/research/cancer-types/melanoma/melanostrum) (14) were whole-exome sequenced as a part of previously-published study for variant discovery (8). Sequencing was performed using NimbleGen’s SeqCap EZ Human Exome Library v2.0 or v3.0 (Roche NimbleGen, Inc., Madison, WI, USA) and an Illumina HiSeq (Illumina, Inc., San Diego, CA, USA); sequencing, alignment and variant calling was performed as described previously (8). Variants and population frequencies were annotated using ANNOVAR (21) and InterVar (python version 2.1.2 20180603) (22). US melanoma families were exome-sequenced in the same manner, while Australian families were exome-sequenced as previously described (23). This overall study was approved by IRB at the National Cancer Institute, as well as local institutions. All subjects provided written informed consent.

### Variant confirmation and genotyping in the population of the same country as the proband

The presence of *NRAS* c.170A>C (p.D57A) in family members was confirmed via Sanger sequencing using primers (F:5’-ACACCCCCAGGATTCTTACAG-3’; R: 5’-GATTCAGAACACAAAGATCATCC-3’) flanking the A170C region of *NRAS* coding sequences. Genomic DNA of the family members were generated from whole blood. Sequencing was performed on an ABI 3730xs DNA sequencer and sequence traces were analyzed using Sequencher. Frequency of *NRAS* c.170A>C (p.D57A) in 6 154 melanoma cases and 2 917 healthy controls from the Melanostrum Consortium (13, 14) (**S1 Table**) was assessed using a custom TaqMan genotyping assay with DNAs from two heterozygous carriers on every assay plate as positive controls. Genotyping was performed using the following primers and probes (primers: F:5’-TCTCTCATGGCACTGTACTCTTCT-3’, R:5’-TGGTTATAGATGGTGAAACCTGTTTGT; probes: 5’-FAM-CCAGCTGTAGCCAGTAT-3’, 5’-VIC-TGTCCAGCTGTATCCAGTAT-3’). Genotyping was performed using 5 uL reaction volumes consisting of: 2.5 uL of 2X KAPA Probe Fast MasterMix (Kapa Biosystems, Woburn, MA), 0.25 uL of 20X TaqMan® assay-specific mix of primers and probes, and 2.25 uL of MBG Water. PCR was performed using 9700 Thermal Cycler (ThermoFisher, Waltham, MA, USA) with the following conditions: 95°C hold for 3 min, 40 cycles of 95°C for 3 sec and 62°C for 30 sec, and 10°C hold. Endpoint reads were evaluated using the 7900HT Sequence Detection System (ThermoFisher). We also assessed the frequency of *NRAS* c.170A>C (p.D57A) using additional germline exome sequencing data (sequenced as described above) from 1 371 lung cancer cases and 831 healthy controls from the Environment and Genetics in Lung Cancer Etiology study (EAGLE; https://eagle.cancer.gov) (15).

### NRAS expression constructs

An *NRAS* cDNA clone was purchased from Origene (Cat# RC202681), the C-terminal Myc-DDK tag was removed and replaced with a stop codon immediately following the NRAS coding sequence via PCR. p.G12V (c. 35G>T) and p.D57A (c. 170A>C) mutations were introduced using a PCR-based site-directed mutagenesis protocol as described (Agilent Technologies, Cat #200522-5). The sequences of all constructs were verified via Sanger sequencing.

#### Cell culture, transfection, EGF stimulation, GST pull down, and western blotting

COS-7 (ATCC CRL-1651) and 293FT (ThermoFisher Scientific R7007) cells were cultured in DMEM medium (Quality Biologicals) containing 10% FBS. Where indicated, cells were treated with recombinant human EGF protein (R&D, Cat# 236-EG-200) in medium free of FBS. *NRAS* constructs (wild-type, p.G12V (c.G35T), and p.D57A (c.A170C) were transfected into COS7 cells using PolyFect Transfection reagent (Qiagen, Cat# 301105) while the transfection of 293FT cells was performed using Lipofectamine 2000 from Invitrogen. Two days after transfection, total cell lysates were generated in RIPA buffer (Thermo Scientific) and subjected to water bath sonication (Bioruptor) Samples were resolved in NuPage 4-12% Bis-Tris gel (Invitrogen) electrophoresis. The primary antibodies used were mouse anti-β-actin (Sigma, A5316), mouse anti-Pan AKT (cell signaling, 2920S), rabbit anti-Phospho AKT(S473) (cell signaling, 4060S), rabbit anti-total ERK1/2 (cell signaling, 9106S), mouse anti-Phospho ERK1/2 (Cell signaling, 4695S), and mouse anti-RAS (Millipore Cat# 05-516). To determine GTP-bound NRAS level in cells expressing WT or mutant NRAS proteins, RAS-GTP was pulled down by RAF1-RBD conjugated agarose beads followed by western by anti-RAS antibody (Millipore, Cat17-218).

### Molecular modeling

The coordinates of wild-type GTP-bound NRAS were generated based on the structure of H-RAS with residue modifications (24). The wild-type GTP-bound NRAS was mutated at D57A to understand the mutational effect. The wild-type and GTP-bound NRAS D57A proteins were solvated in the explicit-solvent solvent box of ∼69*69*69 Å^3^. Ions (Na^+^ and Cl^-^) were used to neutralize and generate the 0.15M ionic strength in the system. Molecular dynamics (MD) simulations were performed by the NAMD package with the updated CHARMM all-atom additive force field (version C36) (25, 26). The simulations used the NPT (constant number of atoms, pressure, and temperature) ensemble at a temperature of 310 K and pressure of 1 atm. Short-range van der Waals (VdW) interactions were calculated by the switch function and the long-range electrostatic interactions were calculated by Particle mesh Ewald (PME) method. 500 ns simulations were individually performed for wild-type and mutated NRAS with a time step of 2 fs. Analysis was performed using CHARMM, VMD tools and scripts.

## Data Availability

The data that support the findings will be available in dbGaP at https://www.ncbi.nlm.nih.gov/gap/ following a 6-month embargo from the date of publication of the article in a peer-reviewed journal.

## Acknowledgements

This research was supported [in part] by the Intramural Research Program of the NIH, National Cancer Institute, Division of Cancer Epidemiology and Genetics (ZIACP010201 for KMB, ZIACP101231 for MTL, and ZIACP010144 for MS; https://dceg.cancer.gov/) and Center for Cancer Research (ZIABC010442 for RN; https://ccr.cancer.gov/) and the National Health and Medical Research Council of Australia (1117663 for NKH; https://www.nhmrc.gov.au). This project has been funded in whole or in part with federal funds from the National Cancer Institute, National Institutes of Health, under contract HHSN26120080001E. The content of this publication does not necessarily reflect the views or policies of the Department of Health and Human Services, nor does mention of trade names, commercial products, or organizations imply endorsement by the U.S. Government. Funders had no role in study design, data collection and analysis, decision to publish, or preparation of the manuscript. All simulations were performed using the high-performance computational facilities of the Biowulf PC/Linux cluster at the National Institutes of Health, Bethesda, MD (https://hpc.nih.gov/). Membership of the Melanostrum Consortium can be found at the following link: https://dceg.cancer.gov/research/cancer-types/melanoma/melanostrum

## Supplementary information

**S1 Fig. Pedigree of the extended family harboring *NRAS***^**D57A**^ **annotated with sequencing and genotyping results**. Individuals diagnosed with melanoma, other cancers, and dysplastic nevi are annotated, and the proband is noted with an arrow.

**S1 Table. MelaNostrum Consortium samples genotyped via TaqMan for *NRAS* p.D57A/c.A170C**.

