## Supplementary information for "Novel MAPK/AKT-impairing germline *NRAS* variant identified in a melanoma-prone family"

**S1 Fig**. **Pedigree of the extended family harboring *NRAS*$^{D57A}$ annotated with sequencing and genotyping results.** Individuals diagnosed with melanoma, other cancers, and dysplastic nevi are annotated, and the proband is noted with an arrow.

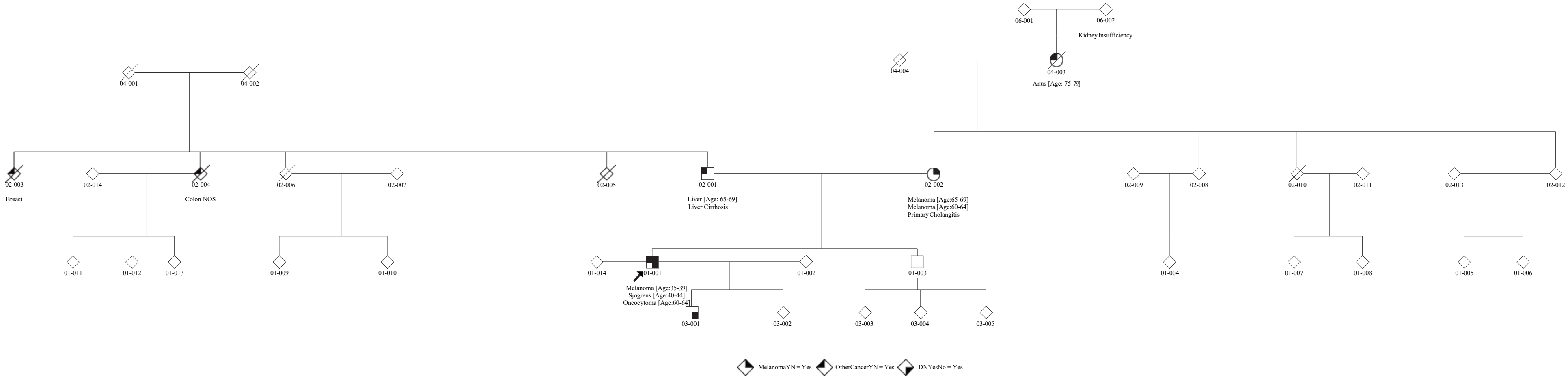

**S2 Table**. MelaNostrum Consortium samples genotyped via TaqMan for *NRAS* p.D57A/c.A170C

| **Sample Origin** | **Case** | **Control** | **Total** |
|---|---|---|---|
| Cyprus | 28 | 157 | 185 |
| Greece (Athens) | 1085 | 510 | 1595 |
| Italy | | | |
| *Cesena* | 247 | 379 | 626 |
| *Genoa* | 587 | 177 | 764 |
| *L'Aquila* | 482 | 67 | 549 |
| *Milan* | 97 | 142 | 239 |
| *Padua* | 1 | 5 | 6 |
| *Rome* | 129 | | 129 |
| *Seveso* | | 279 | 279 |
| Spain | | | |
| *Barcelona* | 2110 | 234 | 2344 |
| *Valencia* | 1388 | 967 | 2355 |
| **Total** | **6154** | **2917** | **9071** |
